# Multimodal latent composites are associated with cognition and Alzheimer’s disease dementia: a framework for systems-level brain health

**DOI:** 10.64898/2026.02.21.26346745

**Authors:** Ella Rowsthorn, Ying Xia, Michael Breakspear, Jurgen Fripp, Gail A. Robinson, Nicholas Ashton, Henrik Zetterberg, Michelle K. Lupton, Meng Law, Matthew P. Pase, Ian H. Harding

## Abstract

Biomarkers from diverse methodological domains are increasingly important in the detection, diagnosis and tracking of neurological diseases and brain health, yet they are often evaluated in isolation. Statistical integration approaches, such as factor analysis, provide a means to combine complementary biomarkers and capture higher-order domains of brain health. Exploratory factor analysis has previously been employed to identify latent brain health constructs using multimodal MRI, fluid biomarkers and cardiovascular risk factors in a non-clinical older population. The current study aimed to validate this integrative framework using confirmatory factor analysis in an independent cohort and test construct associations with cognition and diagnosis of mild cognitive impairment (MCI) or Alzheimer’s disease (AD) dementia.

Data were analysed from 197 participants in the Prospective Imaging Study of Ageing, including 157 cognitively normal controls (CN), 18 participants with MCI and 22 participants with early AD dementia. MRI, cardiovascular, and plasma biomarker processing closely replicated previous methods. Confirmatory factor analysis was conducted in CNs to validate the previously reported latent constructs. Weighted factor composites were then compared between each diagnostic group and tested for associations with cognitive performance (verbal reasoning, verbal memory, visual memory and language) and sensitivity to MCI and AD diagnosis.

Three factors were reproducible across cohorts: 1) Brain & Vascular Health (hippocampal and ventricular volumes, cerebral blood flow); 2) White Matter (WM) Fluid Dysregulation (Free Water, WM enlarged perivascular spaces); and 3) Blood Biomarkers (GFAP, NfL, pTau181). Compared to the CN group, both MCI (β=−1.25, SE=0.19, *p*<.001) and AD dementia (β=−1.52, SE=0.16, *p*<.001) participants had lower Brain & Vascular Health composite scores. MCI (β=0.80, SE=0.20, *p*<.001) and AD dementia (β=1.85, SE=0.17, *p*<.001) participants also had higher Blood Biomarkers composite scores than CNs, but there was no difference in WM Fluid Dysregulation scores across groups (F_(2,192)_= 0.89, *p*=.411). The Brain & Vascular Health composite had the strongest association with MCI/AD dementia among all individual measures and composites. Across all participants, Brain & Vascular Health and Blood Biomarkers composite scores were associated with tests of cognition (*p*<.0125), while WM Fluid Dysregulation did not show any significant associations.

These findings demonstrate that reproducible, multimodal composites can index distinct yet complementary dimensions of brain health relevant to cognition and AD dementia. Importantly, this work highlights the value of an adaptable, integrative framework for combining imaging and plasma biomarkers to characterise system-level brain health and support early detection and mechanistic investigation of cognitive decline and neurodegenerative disease.

## Introduction

Biomarkers of brain health are increasingly central to the diagnosis, monitoring and risk stratification of neurodegenerative disease, often spanning multiple biological systems and measurement modalities, including MRI blood-based assays and clinical assessments^1,2^. This diversity of biomarkers provides the ability to capture complementary aspects of brain health across multiple biological scales. Structural, diffusion- and perfusion-derived MRI metrics provide non-invasive indices of tissue integrity, microvascular function and fluid-regulatory processes – such as cortical thickness, vascular lesions, and extracellular fluid accumulation – that capture complementary features relevant to brain aging and neurodegenerative progression^3–7^. Plasma biomarkers, including phosphorylated tau (pTau), neurofilament light (NfL) and glial fibrillary acidic protein (GFAP), reflect molecular processes linked to Alzheimer’s disease (AD), neuroinflammation and neuronal injury^8^, while cardiometabolic indicators such as cholesterol profiles and blood pressure provide additional insight into vascular health risks and resilience^9,10^. Together, these measures reflect interacting biological processes that synergistically contribute to brain health and vulnerability to AD, vascular cognitive impairment and mixed dementia.

Despite their complementary nature, in practice these biomarkers are often examined in isolation. For example, MRI measures of hippocampal atrophy in AD^11,12^, diffusion metrics of white matter integrity in small vessel disease^13^, and plasma protein levels relevant to AD and brain health in aging populations^14–16^ are commonly reported in isolation. This siloed approach limits interpretability and overlooks the information embedded in the shared covariance across measures, which may offer insights into overarching biological systems, mechanisms, and interacting domains of brain health. Multivariate, multimodal approaches, address this limitation by integrating biomarkers^17^, providing a principled means of developing a broader and richer understanding of brain health.

Recently, we applied exploratory factor analysis to multimodal MRI, cardiovascular, and biofluid biomarkers in adults aged 55 to 80 years from the community^18^. We identified distinct latent constructs that reflected distinguishable aspects of brain health, including clusters of metrics related to brain morphology and neurovascular health, macro- and micro-structural integrity, fluid transport dysregulation, plasma protein biomarkers of neurodegenerative disease, and plasma biomarkers of neuronal injury^18^. Across individuals, weighted composites derived from the factor loadings of these constructs were largely associated with age, but few were inter-correlated, suggesting that these constructs represent complementary dimensions of brain health. However, external validation is necessary to establish generalisability beyond the discovery dataset and it remains unclear whether these constructs are sensitive to variability across the cognitive continuum, including in people with MCI and dementia.

AD is the leading cause of dementia, and brain changes due to AD can precede clinical impairment by up to two decades^19,20^. Although pathological amyloid-β (Aβ) and tau are central pathological hallmarks of AD, converging evidence highlights broader, system-level disturbances in brain structure and physiology that contribute to dementia risk and cognitive decline. These include breakdown of the neurovascular unit^21,22^, impaired clearance of interstitial fluid^23–25^, microstructural degeneration^26,27^, neuroinflammation and glial activation^28^, and elevation of other pathological proteins such as α-synuclein and TAR DNA-binding protein (TDP)-43^29^, which may independently or synergistically increase vulnerability to cognitive impairment and accelerate clinical progression. AD dementia can therefore be conceptualised as a multi-faceted disorder, necessitating biomarkers that probe distinct but interacting domains of brain health. Within this systems-level framework, MCI and early AD dementia provide a valuable test cases for assessing whether latent constructs of brain health are sensitive to cognition and neurodegenerative disease.

The present study evaluated the validity and utility of our previously derived multimodal constructs in an independent cohort^18^. Specifically, our aims were to: (i) confirm the latent construct structure via confirmatory factor analysis in cognitively normal controls (CN) and derive CN-anchored composite weights; (ii) assess differences in composite scores across CN, MCI, and AD dementia groups; and (iii) test the strength of association between the composites and diagnostic status relative to conventional individual biomarkers. By positioning these composites alongside established biomarkers, we aimed to determine if latent dimensions of brain health derived from this integrative framework are replicable and sensitive to cognitive impairment and AD dementia.

## Materials and methods

### Study participants

Cross-sectional data from the Prospective Imaging Study of Ageing (PISA) study was used for the present analysis. The sample included cognitively normal controls (CN) and participants with mild cognitive impairment (MCI) or AD dementia who had complete MRI, MRI/PET and plasma biomarker data.

CN participants were 40-70 years of age and were drawn from the ‘PISA Online’ study, which was originally recruited from existing local genome-wide association cohorts. A subset of participants were twins (5% monozygotic and 10% dizygotic)^30^. Selection was designed to enrich the sample for individuals at extremes of AD genetic risk, based on AD polygenic risk scores and *APOE* ε4 carrier status, in order to maximise power to detect biological and/or cognitive differences associated with future AD risk, as previously reported^30^. All CN participants did not meet diagnostic criteria for MCI or AD (see below).

Separately, participants with MCI and AD dementia were referred by local clinicians following clinical assessment suggestive of AD dementia or a phenotype associated with elevated risk of progression to AD dementia (e.g., amnestic MCI), as previously described^30^. Referred individuals were eligible if they could provide informed consent (with a witness/informant where appropriate); met DSM-5 criteria for an Alzheimer’s-type cognitive disorder^31^ or NIA-AA criteria for MCI or Alzheimer’s-type dementia^32^; scored >20 on the Mini-Mental State Examination (MMSE); and had a Clinical Dementia Rating (CDR) of 0.5 or 1.0. Final diagnostic classification of MCI or AD dementia was confirmed through multidisciplinary review of a structured interview assessing activities of daily living (ADL) and neuropsychological assessments, with AD pathology further confirmed by PET amyloid imaging, as described previously^33^.

For all participants, exclusion criteria included significant neurological conditions other than AD dementia (including history of stroke, diagnosis of mixed dementia, other dementia subtypes, or other neurodegenerative diseases); significant medical condition or history of psychiatric illness that may affect neuropsychological testing (e.g., chronic renal failure, major depression with psychotic symptoms, bipolar disorder); history of neurosurgery; history of alcohol or substance abuse; and contraindication to MRI.

### MRI acquisition

All participants underwent an MRI scan and a separate MR/PET scan, typically within three months of the MRI (median=3.02 months, Q1=2.53, Q3=3.72). Briefly, MRI scans were acquired on a 3T Siemens Prisma with a 64-channel head coil, including: T2-weighted fluid attenuation inversion recovery (FLAIR; TR/TE = 5000/388ms, 1.0mm isotropic resolution), and multi-shell diffusion-weighted imaging (DWI; TR/TE = 4700/84ms, 2.0mm isotropic resolution, 12 x b=0s/mm^2^, 20 x b=1000s/mm^2^, 32 x b=2000s/mm^2^, 60 x b=3000s/mm^2^, alternating AP/PA phase encoding). MRI/PET scans were acquired on a 3T Siemens Biograph mMR hybrid scanner 90-minutes after participants were intravenously injected with [18F]-florbetaben, including: T1-weighted MPRAGE (TR/TE = 2.3/2.26ms, 1mm isotropic), and pseudo-continuous arterial spin labelling (pCASL; TR/TE = 5100/13ms, PLD = 1300ms, labelling duration = 1500ms, 3.4 x 3.4 x 4.0mm resolution) scans^30^.

### MRI analysis

MRI processing followed the procedures outlined in our original exploratory factor analysis study as closely as possible^18^. In brief, brain volumetric outcomes and tissue segmentations were derived from T1w images via Freesurfer^34^ (v6.0.1), including hippocampal volume, ventricle volume, cortical thickness, estimated total intracranial volume, and white and gray matter segmentations. To account for head size, hippocampal volume and ventricle volume were expressed as a ratio of intracranial volume.

White matter hyperintensities (WMH) were automatically segmented by MARS-WMH, a deep-learning algorithm that utilises T1w and FLAIR images to identify areas of relative hyperintensities (nnU-Net algorithm v1.0.1)^35,36^. For consistency with processes previously used, segmentations were masked to the cerebral white matter defined by Freesurfer (i.e. excluding WMH identified in the cerebellum or brain stem), finally derived as WMH total volume.

Enlarged perivascular spaces (ePVS) were automatically segmented by PINGU, an nnU-Net trained on heterogenous data to identify ePVS on T1w images (v.1.0)^37^. N4 bias field correction was applied to the T1w images via ANTs prior to segmentation via PINGU. Resulting ePVS segmentations were sectioned into the basal ganglia (BG; including the caudate, putamen, pallidum, accumbens, thalamus and ventral diencephalon) and normal appearing white matter (WM; white matter minus voxels identified as WMH) regions. ePVS volume with the BG and WM regions were divided by their respective regional volumes to calculate BG and WM ePVS volume fraction.

Isotropic diffusion volume fraction, or ‘Free Water’, was derived from the diffusion images via NODDI three-compartmental modelling^38^. Firstly, diffusion images were pre-processed via an adaptation of an in-house pipeline^39^, including denoising^40^, distortion correction, motion correction and removal of intensity inhomogeneities. Pre-processed images were analysed via the NODDI MATLAB toolbox (v1.0.5, MATLAB R2020b). Resulting maps were masked to the white matter region defined by Freesurfer, and an upper threshold of 0.5 was applied to account for artificially elevated signal around areas of CSF related to partial-volume effects^41^.

Free Water-corrected fractional anisotropy (fwFA) was also derived from the pre-processed diffusion data using the bi-tensor Pasternak model^42,43^. Volumes collected with b-values >1900s/mm^2^ were excluded from this analysis as the highly directionally-dependent data violates model assumptions of Gaussian spatial distribution^44^. The remaining pre-processed images were analysed by the Pasternak model to quantify fwFA^42,43^.

Cerebral blood flow (CBF) was analysed from pCASL images via FSL BASIL^45^ (v4.0.29), which included pre-processing steps for calibration to the M0 image and corrections for distortion, motion and partial volume effects, accompanied by tissue delineations from fsl_anat (FSL v6.0.7.16).

After processing, WMH segmentations and Free Water, fwFA and CBF maps were linearly co-registered to the T1w space via ANTs^46^ (v2.5.3) (or FLIRT via BASIL for CBF), using a nearest neighbour interpolation for WMH segmentations. Using Freesurfer segmentations of white matter and gray matter, we derived total WMH volume, mean white matter Free Water, mean white matter fwFA and mean gray matter CBF.

Raw images and derived outcomes underwent a combination of quantitative and qualitative quality control, where outcomes were excluded if they were likely to be affected by scan quality or processing inaccuracies. Briefly, all T1w and FLAIR images were processed through MRI QC^47^ (v23.1.0), where scans with signal-to-noise ratio, contrast-to-noise ratio, entropy-focus criterion (EFC), foreground-to-background energy ratio (FBER) and voxel smoothness (FWHM) outside the inter-quartile range for the cohort were flagged for review. For segmentation tasks, including for WMH and ePVS, data were excluded if motion artifacts impacted segmentation accuracy or poor scan quality impaired differentiation of features. Visual inspection of raw diffusion data was performed to ensure complete brain coverage and to identify severe head motion or other significant image artefacts. CBF maps were inspected for noisy signal in areas typically susceptible to artifact (such as near the sinuses), and mean values were assessed for outliers relative to the cohort and physiological range. All co-registrations to T1w space were inspected for accuracy.

### Fluid biomarkers

Blood samples were collected from all participants, typically on the same day as the MRI scan (or within one month of other assessments such as MRI/PET). Approximately half of the samples were taken after an overnight fast (55%). As previously described, high-density lipoprotein (HDL) and low-density lipoprotein (LDL) cholesterol were quantified, and *APOE* genotyping was performed^30,48^. Participants were categorised as an *APOE* ɛ4 allele carrier (either homozygote or heterozygote) or non-carrier.

Plasma samples were aliquoted and stored a -80°C freezer prior to analysis. Plasma Aβ40, Aβ42, GFAP and NfL were analysed with the Neurology 4-Plex Assay (Quanterix, Billeric, MA, USA). Coefficients of variance, estimated using quality control samples with clinically relevant biomarker concentrations, were below 10%.

### Cardiovascular risk indices

Measurement of office blood pressure, height and weight were completed in-person by trained researchers on the same day as the MRI scan. Body mass index (BMI) was calculated as weight/height^2^. Seated blood pressure was measured in mmHg, after at least five minutes of seated rest, via an OMRON HEM-907 device.

### Neuropsychology

Neuropsychological testing was administered in person by trained neuropsychologists, spanning several domains of cognition. Tests included: Similarities^49^ to assess verbal reasoning/abstraction; the Rey Auditory Verbal Learning Test (RAVLT) immediate recall^50^ to assess verbal memory; the Topographical Recognition Memory Test (TRMT)^51^ to assess visual memory; and the FAS^52^ to assess verbal fluency. For CNs, neuropsychological testing was typically administered on the same day as MRI (92%, with the longest time interval being one month). For participants with MCI or AD dementia, neuropsychological testing was completed within eight months of MRI (median=2.17 months, Q1=0.91, Q3=2.68).

### Statistical analysis

#### Exploratory factor analysis

Firstly, we set out to validate a previously defined and reported factor structure from the Brain and Cognitive Health (BACH) cohort study which enrolled adults from the general community (N=127: aged 55-80; mean age = 67 years; 68% women; previously reported)^18^. The plasma marker pTau217 included in the original BACH analysis was not available in the PISA dataset. To allow for confirmatory factor analysis in PISA, which requires complete data correspondence, we re-conducted the exploratory factor analysis in the BACH cohort after excluding pTau217.

#### Confirmatory factor analysis

We then evaluated the reliability of the factors structure in controls from the PISA cohort using Bayesian confirmatory factor analysis via the ‘blavaan’ package^53^ (version 0.5-9) in R. A Bayesian approach was adopted to improve parameter stability and address estimation issues observed under maximum likelihood estimation, including a standardised factor loading marginally exceeding 1.0 (i.e. a Heywood-type case). Bayesian estimation applies weakly informative priors that induce shrinkage toward plausible parameter ranges, thereby reducing the influence of sampling variability and improving the robustness of factor loading estimates. All CN participants were included in the factor analysis, and all observed variables were z-scored to ensure scaling was consistent across measures. As cross-loadings are not permissible in confirmatory factor analysis, BG ePVS was removed from the Brain & Vascular Health factor and retained on the Structural Integrity to maintain factor interpretability and adequate size. Model adequacy was evaluated using posterior predictive p-values, where values closer to 0.50 indicate excellent fit, and values <0.05 or >0.95 indicate a poor fit. Factor loadings with low posterior support (standardised loadings less than 0.30) were removed iteratively, guided by model fit.

After identifying replicable factors, we generated an individual-level composite score for each factor to enable subsequent analysis with clinical outcomes. For each participant, weighted composites were calculated relative to the CN means using the variable loadings estimated during the confirmatory factor analysis, as per the equation below:

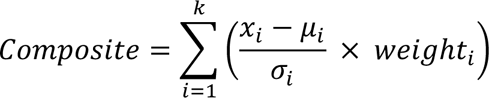

Here, *k* is the number of variables in a given factor, *μ_i_* and *σ_i_* are the CN group’s mean and standard deviation for variable *i*, and *weight* refers to corresponding factor loading. This approach yields standardized composites that reflect the original factor structure and allows scores for individuals with MCI or AD dementia to be interpreted relative to the CN distribution. CN group means used in scoring are provided in **Table 2**.

#### Associations with clinical diagnosis

We examined whether the latent composite scores from each factor differed significantly across diagnostic groups (CN, MCI and AD dementia) using ANCOVA models, adjusting for age and sex. Where the overall group effect was statistically significant, we conducted post hoc pairwise comparisons of estimated marginal means with Bonferroni correction to examine differences between each group (i.e., CN vs MCI, CN vs AD dementia, MCI vs AD dementia). For the Blood Biomarker composite, we conducted a sensitivity analysis by including fasting status as an additional covariate. Notably, although the MCI (n=18) and AD (n=22) group sizes are typical for clinical neuroimaging studies, they were much smaller than the CN group (n=157). To assess the potential impact of this imbalance, we conducted a further sensitivity analysis by repeating the ANCOVA tests using a subset of 40 CN participants propensity-matched for age and sex via the ‘matchit’ software^54^ in R (v4.7.2; see **Supplement** for details and subset summary).

We conducted binomial logistic regressions to estimate and rank the odds ratios of each composite score for association with MCI/AD, adjusting for age and sex. For comparison, we also examined all individual variables included in the original exploratory factor analysis: HDL and LDL cholesterol levels, WMH volume (log +1 transformed), systolic blood pressure and plasma Aβ42/40. PET amyloid centiloid was additionally included as a well-established marker of AD pathology^55^.

#### Associations with cognition

Finally, across the whole cohort, we investigated whether the composite scores were associated with cognitive performance using multiple linear regressions adjusted for age, sex, and years of education. Two CN participants did not have cognitive data available and were not included in analyses relating to cognition. We adjusted for multiple comparisons across the four cognitive tests (Similarities, RAVLT, TMRT and FAS) with Bonferroni correction, where a *p*-value of ≤ .0125 was considered statistically significant.

## Results

### Exploratory factor analysis without pTau217

We firstly evaluated whether the previously reported five-factor solution remained appropriate for the data without inclusion of pTau217 in the BACH cohort. Scree plot inspection (**eFigure 1**) suggest that either a four-factor or five-factor solution could fit the data. Although the five-factor model showed excellent fit indices (root mean square error of approximation = 0.03, Tucker-Lewis index = 0.96), the “AD Biomarkers” factor was reduced to a single indicator, Aβ42/40, with a loading >1.00, indicating overfitting (**eTable 1**). The four-factor model demonstrated very good fit (root mean square error of approximation = 0.04, Tucker-Lewis index = 0.93), preserved a similar organisational structure and did not have a single-indicator factor. Therefore, we adopted the four-factor solution. Variables with loadings >0.30 were considered to meaningfully contribute to each factor. Final loadings closely matched our previous analysis^18^, with the exception that pTau181 now clustered with GFAP and NfL, forming a broader “Blood Biomarkers” construct (**Table 1**).

**Table 1.** Four factors were identified in the exploratory factor analysis without pTau217.

|  | <u>Factor 1:</u><br>Brain & Vascular<br>Health | <u>Factor 2:</u><br>Structural Integrity | <u>Factor 3:</u><br>WM Fluid<br>Dysregulation | <u>Factor 4:</u><br>Blood Biomarkers |
| --- | --- | --- | --- | --- |
| HDL Cholesterol | 0.41 | -0.03 | 0.21 | 0.18 |
| BMI | -0.41 | 0.14 | 0.05 | -0.25 |
| Hippocampal Volume | 0.82 | -0.07 | -0.06 | -0.04 |
| Ventricle Volume | -0.70 | -0.06 | -0.11 | -0.02 |
| CBF | 0.41 | 0.11 | -0.03 | 0.03 |
| BG ePVS | 0.40 | 0.36 | 0.13 | -0.14 |
| fwFA | -0.09 | 0.63 | -0.08 | 0.24 |
| Cortical Thickness | 0.03 | 0.59 | 0.12 | -0.16 |
| WM ePVS | 0.05 | 0.03 | 0.98 | 0.02 |
| Free Water | -0.26 | -0.12 | 0.56 | 0.17 |
| pTau181 | -0.06 | 0.09 | 0.06 | 0.32 |
| GFAP | 0.08 | 0.03 | 0.05 | 0.64 |
| NfL | -0.04 | 0.03 | -0.02 | 0.66 |

**Note:** Loadings for each variable are reported, where variables with a loading of 0.30 or greater were considered to meaningfully contribute to the latent factor.
Aβ=amyloid-beta; BG=basal ganglia; BMI=body mass index; CBF=cerebral blood flow; ePVS=enlarged perivascular space; FW=Free Water; fwFA=Free Water-corrected fractional anisotropy; GFAP=glial fibrillary acidic protein; HDL=high-density lipoprotein; NfL=neurofilament light chain; pTau=phosphorylated tau; WM=white matter.

### Independent confirmatory factor analysis

A cohort of 157 CN participants from the PISA cohort was available with complete MRI, plasma biomarker, and cardiovascular health data (mean age was 62 years, 80% were women; **Table 2**). The initial confirmatory factor analysis with all hypothesised factors demonstrated poor model fit (posterior predictive p-value = 0.002), with some indicators showing weak or non-significant loadings, including HDL cholesterol, BMI and fwFA (**eFigure2**). After iteratively trimming low-loading (<0.30) variables (removing HDL cholesterol, BMI and fwFA), four robust factors remained, showing acceptable model fit (posterior predictive p-value = 0.285). However, BG ePVS volume fraction had an inverse direction within the Structural Integrity factor than previously defined. To better understand this contradictory finding, we conducted a post-hoc linear regression investigating the relationship between BG ePVS and age, reported in the **Supplement**. Unlike in the original factor analysis cohort, BG ePVS had a *positive* association with age in the present cohort (**eFigure 3**), suggesting BG ePVS volume may be a conceptually inconsistent or contextually dependent marker of brain health or aging (see **Discussion**). As theoretical consistency is an important element in confirming the validity and replicability of these constructs, we did not proceed with the “Structural Integrity” factor. Thus, the final three validated factors were: “Brain & Vascular Health” comprised of greater hippocampal volume and CBF, and lower ventricle volume; “WM Fluid Dysregulation” comprised of greater Free Water and WM ePVS; and “Blood Biomarkers” comprised of greater GFAP, NfL and pTau181 (**Table 3**).

**Table 2.** PISA Cohort Summary.

|  | **CN**<br>n=157 | **MCI**<br>n=18 | **AD**<br>n=22 |
| --- | --- | --- | --- |
| **Demographics** |  |  |  |
| Age at MRI, years | 61.68 (6.72) | 69.31 (6.89)*** | 65.47 (6.94)** |
| Women (n, %) | 125 (79.6%) | 7 (38.9%)*** | 13 (59.1%) |
| Education, years | 13.5 (2.76) | 12.61 (3.62) | 14.18 (3.40) |
| *APOE* ε4 carrier (n, %) | 91 (58.0%) | 9 (50.0%) | 14 (63.6%) |
| **Cognition** |  |  |  |
| Similarities, total score^ | 32.54 (4.48) | 26.94 (4.99) | 23.95 (8.94) |
| RAVLT, immediate recall score^ | 13.68 (1.46) | 9.61 (3.09) | 11.00 (3.24) |
| TMRT, total score^ | 25.95 (2.84) | 22.00 (4.99) | 17.55 (6.02) |
| FAS, total score^ | 42.23 (11.55) | 35.83 (13.05) | 27.09 (9.76) |
| **Cardiovascular Risk Indices** |  |  |  |
| BMI, index^ | 26.74 (5.25) | 26.40 (3.97) | 25.64 (3.93) |
| Systolic Blood Pressure, mmHg^ | 135 (18) | 137 (18) | 141 (15) |
| HDL Cholesterol, mmol/L^ | 1.68 (0.41) | 1.51 (0.47) | 1.70 (0.47) |
| LDL Cholesterol, mmol/L^ | 3.33 (0.95) | 2.89 (1.05) | 3.56 (0.88) |
| **Neuroimaging** |  |  |  |
| WM Free Water, volume fraction | 0.085 (0.013) | 0.089 (0.014) | 0.084 (0.017) |
| BG ePVS, fraction of BG region | 0.0044 (0.0020) | 0.0046 (0.0018) | 0.0055 (0.0026) |
| WM ePVS, fraction of NAWM region | 0.0033 (0.0026) | 0.0026 (0.0022) | 0.0022 (0.0023) |
| GM cerebral blood flow, ml/100g/min | 23.28 (4.75) | 19.95 (4.75) | 18.43 (4.27) |
| WMH volume, mm<sup>3</sup> (median, IQR) | 337 (760) | 1017.5 (4437.25) | 561.5 (1438.5) |
| Cortical thickness, mm | 2.40 (0.07) | 2.30 (0.06) | 2.27 (0.09) |
| Ventricle volume, fraction of eTIV | 0.0146 (0.0056) | 0.0283 (0.0144) | 0.294 (0.0111) |
| Hippocampal volume, fraction of eTIV | 0.0057 (0.004) | 0.0046 (0.005) | 0.0048 (0.007) |
| fwFA, degree of anisotropy | 0.507 (0.011) | 0.508 (0.013) | 0.497 (0.035) |
| PET amyloid centiloid (median, IQR)^ | -5.94 (13.50) | 54.56 (70.05) | 99.14 (40.50) |
| PET amyloid centiloid <20 (n, %)^ | 12 (7.6%) | 12 (66.7%) | 22 (100%) |
| **Plasma Biomarkers** |  |  |  |
| Fasted blood sample (n,%) | 98 (62.4%) | 5 (27.8%) | 3 (13.6%) |
| Aβ42/40 ratio | 0.071 (0.015) | 0.069 (0.011) | 0.064 (0.008) |
| GFAP, pg/mL | 66.52 (43.61) | 97.39 (45.88) | 170.81 (54.99) |
| NfL, pg/mL | 13.45 (8.59) | 18.44 (13.30) | 23.73 (7.87) |
| pTau181, pg/mL | 6.10 (5.31) | 20.53 (18.06) | 25.80 (20.37) |

**Note:** Reported as mean (SD) reported unless otherwise specified.
*APOE* ε4 carrier defined as having at least one ε4 allele (i.e. either heterozygote or homozygote).
^Some outcomes were not available for some participants:
PET amyloid centiloid: total N=188: CN 150, MCI 18, AD 20.
Cognition outcomes: total N=195: CN 155, MCI 18, AD 22.
BMI: total N=179: CN 157, MCI 11, AD 11.
Systolic BP: total N=176: CN 157, MCI 9, AD 10.
HDL and LDL Cholesterol: total N=196: CN 157, MC 18, AD 21
\* $p < .05$ , \*\*\* $p < .001$ : significant difference compared to the CN group in Wilcoxon t-test or Fisher's Chi-Squared test (demographics only)
Aβ =amyloid-beta; AD=Alzheimer's disease; *APOE*=apolipoprotein-E; BG=basal ganglia; BMI=body mass index; CBF=cerebral blood flow; CN=cognitively normal control; ePVS=enlarged perivascular space volume; fwFA=Free Water-corrected fractional anisotropy; GFAP=glial fibrillary acidic protein; HDL=high-density lipoprotein; LDL=low-density lipoprotein; MCI=mild cognitive impairment; NAWM=normal appearing white matter; NfL=neurofilament light chain; PET=positron emission tomography; pTau=phosphorylated tau; RAVLT=Rey Auditory Verbal Learning Test immediate recall; TMRT=Topographical Recognition Memory Test; WM=white matter; WMH=white matter hyperintensity volume.

**Table 3.** Confirmatory Factor Analysis Loadings.

|  | **Standardised**<br>**Factor Loading** | **95% Credible**<br>**Interval** | **R-hat** |
| --- | --- | --- | --- |
| **Brain & Vascular Health** |  |  |  |
| Hippocampal Volume | 0.316 | [0.148, 0.504] | 1.001 |
| Ventricle Volume | - 0.913 | [-1.101, -0.684] | 1.001 |
| CBF | 0.343 | [0.171, 0.544] | 1.000 |
| **WM Fluid Dysregulation** |  |  |  |
| WM ePVS | 0.774 | [0.467, 1.070] | 1.003 |
| Free Water | 0.726 | [0.452, 1.054] | 1.004 |
| **Blood Biomarkers** |  |  |  |
| GFAP | 0.969 | [0.851, 1.120] | 1.001 |
| NfL | 0.629 | [0.492, 0.797] | 1.000 |
| pTau181 | 0.667 | [0.533, 0.828] | 0.999 |

**Note:** Confirmatory factor analysis loadings for the final structure.
BG=basal ganglia; CBF=cerebral blood flow; ePVS=enlarged perivascular space volume; GFAP=glial fibrillary acidic protein; NfL=neurofilament light chain; WM=white matter.

### Composite score group comparisons

A cohort of 18 participants with confirmed MCI (66% with PET amyloid centiloid > 20, suggestive of early AD pathology) and 22 with confirmed AD dementia (all PET amyloid centiloid > 20, mean = 99) from the PISA cohort were available for analysis. Wilcoxon *t*-tests (for continuous measures) and Fisher’s Chi-Squared tests (for categorical measures) revealed that relative to CNs, the MCI group had a lower proportion of women, while both the MCI and AD groups were older (**Table 2**).

Adjusted ANCOVAs revealed significant differences in the Brain & Vascular Health composite across all groups (F_(2,192)_ = 92.97, *p*<.001). Compared to the CN group, both MCI (β=−1.25, SE=0.19, *p*<.001) and AD dementia (β=−1.52, SE=0.16, *p*<.001) participants had lower Brain & Vascular Health composite scores, but there was no significant difference between the MCI and AD dementia groups (β=−0.27, SE=0.22, *p*=.702; **Figure 1a**). Similarly, there was a significant difference in the Blood Biomarkers composite across all groups (F_(2, 192)_ = 76.02, *p*<.001), and this relationship remained after further adjustment for blood sample fasting status (F_(2, 188)_ = 79.00, *p*<.001). Both MCI (β=0.80, SE=0.20, *p*<.001) and AD dementia (β=1.85, SE=0.17, *p*<.001) participants had higher Blood Biomarkers composite scores than CN, and AD dementia participants also had higher Blood Biomarkers than those with MCI (β=1.05, SE=0.24, *p*<.001; **Figure 1c**). WM Fluid Dysregulation composite scores were not significantly different across groups (F_(2,192)_=0.89, *p*=.411; **Figure 1b**). For descriptive purposes, composite scores are additionally presented across diagnostic groups stratified by PET amyloid status in the Supplement (**eFigure 4**). In sensitivity analyses with the propensity-matched CN group, the differences in Brain & Vascular Health composite scores (F_(2,75)_=37.55, *p*<.001) and Blood Biomarkers composite scores (F_(2,75)_=23.03, *p*<.001) across groups were attenuated but remained statistically significant (**eFigure 5**).

**Figure 1.**
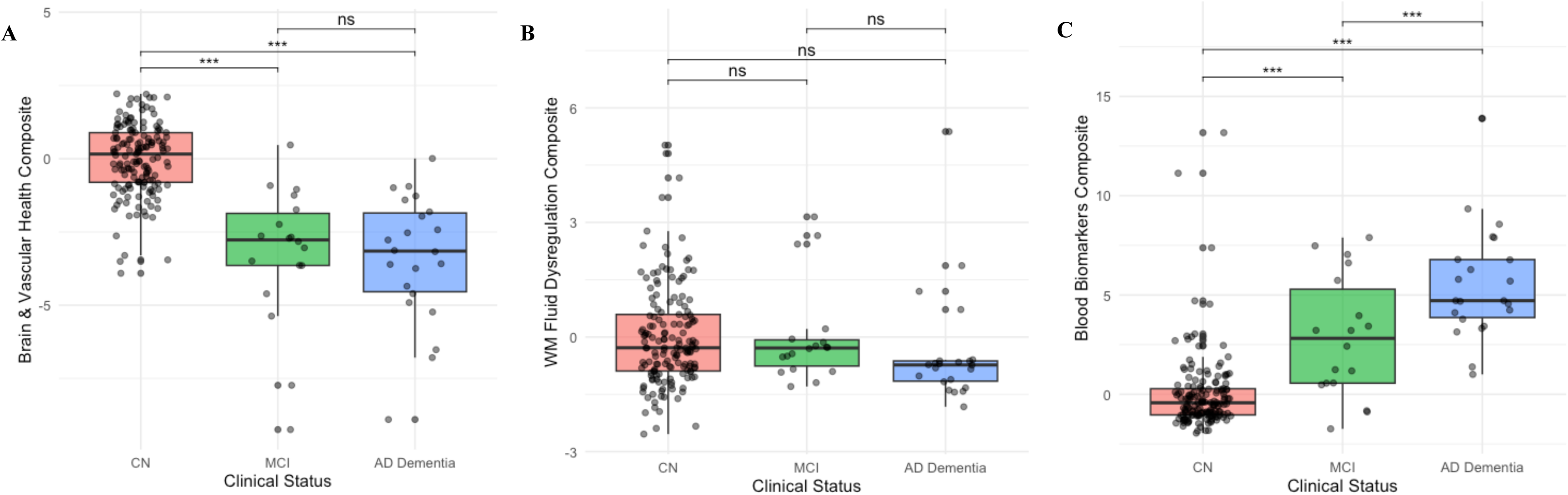
Composite score differences between CN, MCI and AD dementia groups. Differences across cognitively normal controls (CN), mild cognitive impairment (MCI) and Alzheimer’s disease (AD) dementia groups reported for: (**A**) Brain & Vascular Health composite scores; (**B**) White Matter (WM) Fluid Dysregulation composite score; (**C**) Blood Biomarkers composite scores. \*\*\**p*<.001, *ns* = did not meet statistical significance (*p*>.05).

Using logistic regressions, the Brain & Vascular Health composite showed the strongest association with MCI/AD dementia (OR = 0.07, 95% confidence interval [CI] = [0.03, 0.16]), corresponding to an approximately 14-fold reduction in odds for each SD increase in the composite score (**Figure 2**). The Blood Biomarkers composite also had a significant association with the MCI/AD dementia group (OR = 4.68, 95% CI = [2.84, 8.48]). The WM Fluid Dysregulation composite was not significantly associated with the MCI/AD dementia group (OR = 0.69, 95% CI = [0.44, 1.02]).

**Figure 2.**
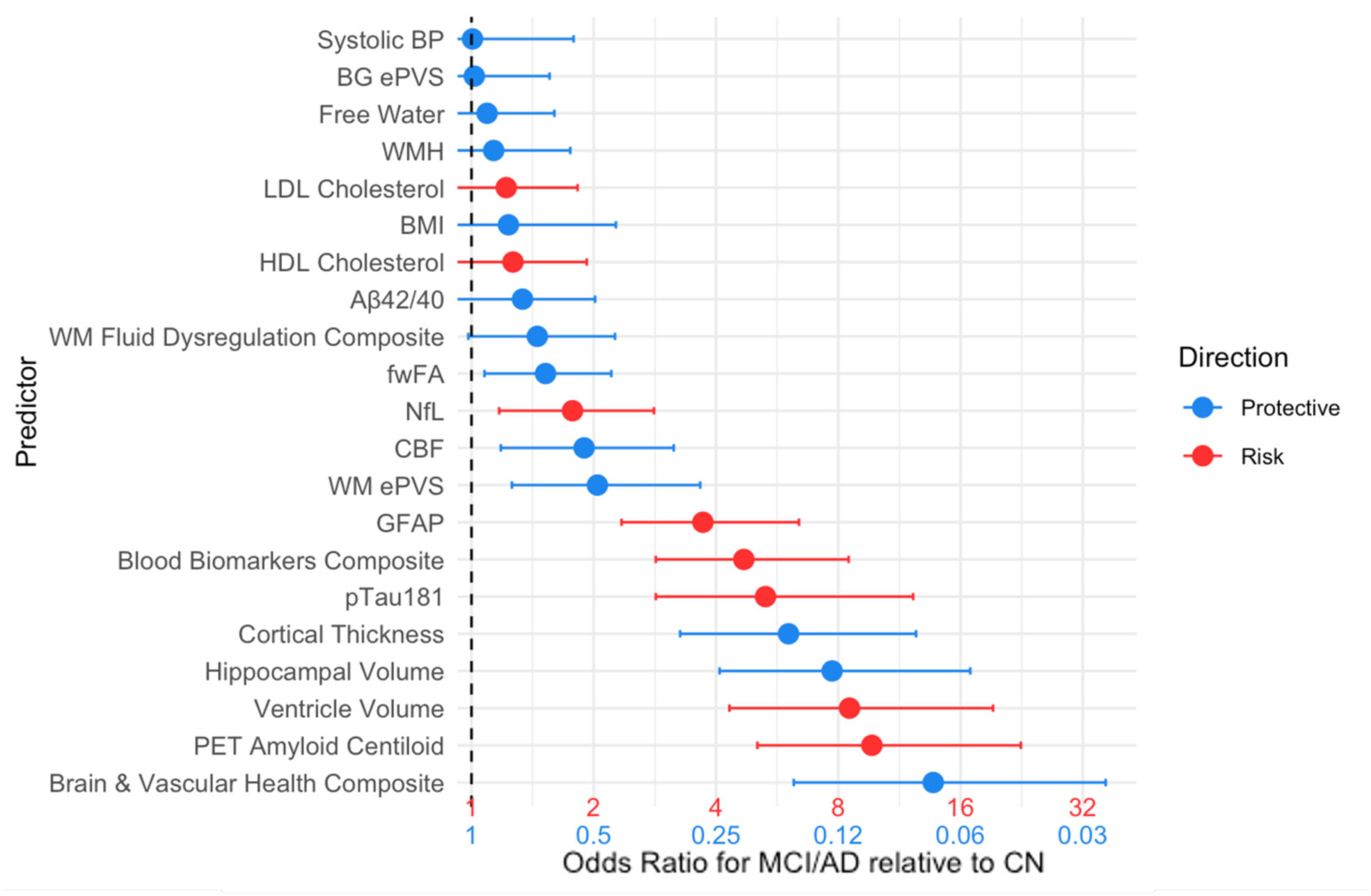
Individual and composite biomarker associations with MCI/AD dementia. Odds ratios (OR) from logistic regression are shown for each predictor, ranked by magnitude. Values reflect the relative odds of MCI/AD dementia as a combined outcome per 1 SD increase in the predictor. Points indicate ORs, and horizontal bars show 95% confidence intervals. For presentation, ORs<1 were inverted so size of effect is displayed in the same direction, instead, ‘protective’ (ORs<1) and ‘risk’ (ORs>1) directions and corresponding x-axis are color-coded as blue and red, respectively. Overlap with the dotted line (OR=1) indicates no significant association with MCI/AD dementia. Aβ=amyloid-beta; AD=Alzheimer’s disease; BG=basal ganglia; BMI=body mass index; CBF=cerebral blood flow; CN=cognitively normal health controls; ePVS=enlarged perivascular space volume; fwFA=Free Water-corrected fractional anisotropy; GFAP=glial fibrillary acidic protein; HDL=high-density lipoprotein; LDL=low-density lipoprotein; MCI=mild cognitive impairment; NfL=neurofilament light chain; PET=positron emission tomography; pTau=phosphorylated tau; WM=white matter; WMH=white matter hyperintensity volume.

### Composite scores and cognition

The Brain & Vascular Health composite was positively associated with each of the cognitive tests, including Similarities, RAVLT, TMRT (β=0.294–0.481, all *p*<.001; **Table 4**), although the association with FAS did not survive FDR-correction (*p*=.015). The Blood Biomarkers composite was negatively associated with all cognitive tests (β=-0.252– -0.426, all *p*<.001; **Table 4**). The WM Fluid Dysregulation composite was not significantly associated with any of the cognitive tests (all *p*>.0125; **Table 4**).

**Table 4.** Composite Score Associations with Cognition.

|  | **Factor 1:**<br>**Brain & Vascular Health** |  | **Factor 2:**<br>**WM Fluid Dysregulation** |  | **Factor 3:**<br>**Blood Biomarkers** |  |
| --- | --- | --- | --- | --- | --- | --- |
| | $\beta$ (SE) | *p* | $\beta$ (SE) | *p* | $\beta$ (SE) | *p* |
| **Similarities** | 0.462 (0.072) | <.001 | -0.010 (0.071) | .885 | -0.310 (0.070) | <.001 |
| **RAVLT** | 0.294 (0.071) | <.001 | 0.028 (0.065) | .669 | -0.266 (0.065) | <.001 |
| **TRMT** | 0.481 (0.073) | <.001 | 0.173 (0.071) | .015 | -0.426 (0.068) | <.001 |
| **FAS** | 0.191 (0.078) | .015 | -0.039 (0.070) | .574 | -0.252 (0.071) | <.001 |

**Note:** Multiple linear regressions between each cognitive outcome and factor constructs. $\beta$ estimates reflect change in cognitive outcome per 1 SD unit in the composite score, adjusting for age, sex and education.
A *p*-value of $\leq .0125$ was considered statistically significant after FDR correction.
RAVLT=Rey Auditory Verbal Learning Test immediate recall; TRMT=Topographical Recognition Memory Test; WM=white matter.

## Discussion

In this study, we provide confirmatory evidence of multimodal latent constructs which are relevant to cognitive health and AD dementia. We externally validated latent constructs previously identified in older adults from the community^18^, of which three demonstrated a consistent and replicable structure in CNs in the present cohort: Brain & Vascular Health (hippocampal volume, ventricular volume, gray matter CBF), WM Fluid Dysregulation (white matter perivascular spaces, extracellular Free Water), and Blood Biomarkers (plasma GFAP, NfL and pTau181). The Brain & Vascular Health and Blood Biomarker composites were associated with cognitive performance and diagnostic status. Notably, Brain & Vascular Health showed the strongest association with MCI/AD dementia out of all individual measures. Importantly, these constructs were first derived and validated in older adults without significant neurological disease or clinical cognitive impairment, and subsequently applied to individuals with MCI or AD dementia. As such, the constructs were not optimised to detect any single clinical syndrome or disease-specific pathology. Rather than being tailored to AD, the constructs reflect broader systems-level processes relevant to brain health. These constructs differentiated cognitively normal individuals from those with cognitive impairment. This underscores the potential value of this approach: a generalisable, disease-agnostic framework for assessing multidimensional brain health in ageing and neurodegenerative disease.

### Composite biomarkers are sensitive to cognition and AD dementia

The validated factors captured distinct constructs of brain health, two of which were sensitive to cognition and AD dementia. We found that the Brain & Vascular Health composite was associated with cognitive performance across multiple domains, including verbal reasoning, verbal memory, visual memory and language. These results align with evidence that neurovascular dysfunction and atrophy represent proximal pathophysiological contributors to cognitive decline and dementia^21^. Notably, the Brain & Vascular Health composite showed a stronger association with diagnostic status than other individual imaging measures (e.g., hippocampal volume or ventricular volume), suggesting that multimodal MRI composites that integrate multiple dimensions of brain structural and vascular integrity may be more sensitive to clinically relevant variation than single markers. The composite also differed significantly between CN and both MCI and AD dementia, indicating sensitivity to brain changes that deviate from cognitively normal-derived means regardless of exact pathology.

Associations between the Blood Biomarker composite, cognition and diagnostic status also aligned with prior research linking GFAP, NfL and pTau181 with neurodegeneration and AD-related processes^56^. Interestingly, pTau181 alone showed a stronger relationship with MCI/AD dementia than the Blood Biomarker composite, which may be attributable to its ability to index amyloid levels as well as tau hyperphosphorylation^56^, offering a more specific measurement of pathology burden. In contrast, GFAP and NfL are likely related to broader, non-specific mechanisms such as astrocyte reactivity and axonal degradation^8^, and are thought to reflect more dynamic aspects of neural damage that often co-vary with active neurodegeneration among other non-neurodegenerative events (e.g., traumatic brain injury, stroke)^57,58^. The Blood Biomarker composite may therefore capture a combination of cumulative pathological burden (indexed by pTau181) and more dynamic injury-related processes (indexed by GFAP and NfL), which could confer greater sensitivity to broad pathology or injury, but not necessarily greater specificity to any disease.

By contrast, we did not observe any significant associations between the WM Fluid Dysregulation composite and either cognition or MCI/AD dementia diagnosis. This finding does not support a direct relationship between impaired interstitial fluid regulation, as indexed by WM ePVS and Free Water, and cognition in this cohort. Interestingly, Free Water showed one of the weakest associations with MCI/AD dementia, while WM ePVS showed an unexpected ‘protective’ association in logistic regression. These opposing effects suggest that their shared variance may reflect processes not uniquely related to fluid system dysfunction, but potentially also attributable to other age-related or compensatory mechanisms. The apparent inverse association of WM ePVS is particularly atypical and may reflect non-linear relationships or mixed pathophysiological pathways, warranting further investigation.

### Validation of factor composites

Confirmatory factor analysis provided external validation and partial confirmation of the factor structure first defined using an exploratory factor analysis in an independent cohort with the same outcome measures. The Brain & Vascular Health construct defined by the confirmatory factor analysis retained hippocampal volume, ventricular volume and CBF, but did not include HDL cholesterol or BMI as per the original exploratory factor analysis. Several non-mutually exclusive explanations could account for this observation. First, HDL cholesterol and BMI are systemic and relatively distal markers of cerebrovascular disease risk. Their relationship with more proximal measures of brain structure and perfusion may be cumulative or mediated by intermediate processes (e.g., endothelial dysfunction, neuroinflammation)^59–61^, where brain health was captured more directly by CBF and brain atrophy measures. Second, cohort differences such as genetic enrichment or small methodological variations (such as in blood sample analysis) could affect covariance patterns and reduce the stability of peripheral markers. *APOE* ɛ4 in particular has a known influence on the relationship between cardiovascular health and brain atrophy^62–64^, and therefore may modify the relationships and factor structure of these markers. As the current sample was underpowered to repeat the factor analysis in *APOE* ɛ4 carrier/non-carrier subgroups, future work should test these possibilities by conducting factor analysis stratified by *APOE* status, alongside replication of cardiovascular risk measures and exploration in longitudinal mediation models capable of distinguishing direct from indirect effects.

The other notable change observed during validation was a reversal in how BG ePVS related to the Structural Integrity factor. In the original exploratory factor analysis, higher cortical thickness, higher fwFA and higher BG ePVS loaded together; however, in the validation confirmatory factor analysis, this factor comprised higher cortical thickness but *lower* BG ePVS. We confirmed that, unlike in the original cohort, BG ePVS was positively associated with age in the validation sample, suggesting that the interpretation of BG ePVS may differ across samples or be unstable. Because this instability impaired factor replicability, we excluded the discordant factor from downstream analyses. Nonetheless, this discrepancy warrants further consideration. One possibility is that BG ePVS reflect biologically heterogenous and context-dependent processes, with their meaning varying across unobserved subtypes or states. For example, BG ePVS may index cerebral small vessel disease^65^, local atrophy and deformation^66,67^, or transient changes in perivascular fluid dynamics^68^, which may also co-occur within individuals and produce non-linear or mixture effects not captured by simple factor models. Consistent with this interpretation, although greater BG ePVS burden is generally considered a marker of poorer brain health, reviews and meta-analyses have highlighted heterogeneity and inconsistency across multiple cohorts^68–70^. Clarifying the role and interpretation of BG ePVS will therefore require approaches that directly link imaging measures to underlying pathology (i.e., biological validation in different contexts).

### Broad application of composite biomarkers

Although this study focused on associations between composite constructs, cognition, and cognitive impairment due to AD, these composites were derived from older adults without known neuropathology (i.e., a ‘normative’ sample) to capture broad domains of brain health rather than disease-specific signatures. Their structure, therefore, reflects typical relationships among measures in older adults, and not necessarily those that emerge in the context of specific neurodegenerative diseases. For example, although hippocampal atrophy and NfL are often correlated in AD^71,72^, they did not load together in the normative factor structure derived from healthy older adults^18^. Applying these normative-reference composites to individuals with cognitive impairment therefore allows assessment of how disease status is associated with deviations in composite scores relative to relationships expected under typical aging, rather than formal differences in factor structure. The underlying factor structure could differ in persons with neurodegenerative disease as compared to CNs, and elucidating such differences may highlight new biological or clinical insights. Nevertheless, a formal exploration of this was not possible given the nature of our sample, and this remains an important future research avenue.

It is also important to emphasise that inclusion of a marker in a given health domain or composite does not suggest its relative importance over other individual markers that did not load onto one of the derived factors. Rather, markers not part of a composite may instead represent outcomes with unique or less shared sources of variance. Individual measures such as WMH, HDL and LDL cholesterol, systolic blood pressure, and plasma Aβ42/40 did not cluster into a latent construct. Nonetheless, plasma Aβ42/40 showed a significant association with MCI/AD in logistic regression models. So, while composites leverage internal similarities across features, stand-alone markers may still provide unique and important biological information. Accordingly, we do not suggest that our composite measures are superior to individual biomarkers. Instead, we provide a framework for probing latent constructs of brain health while potentially reducing noise inherent to individual measures. Their robustness across cohorts supports their potential utility for a broad range of applications, including other neurodegenerative, vascular, traumatic injury or psychiatric conditions where multivariate integration may reveal shared biological pathways^73–75^. Future research should extend these composites to other settings and evaluate their sensitivity across different diseases. Given their broad nature, these constructs may enable comparisons across disease states, support patient stratification in clinical trials using broad indices of brain health, and facilitate monitoring of disease progression within specific domains that are not tied to any single pathology.

While the constructs identified within this study are likely to require further refinement and validation across different populations and disease contexts, harmonisation and normative modelling approaches will be essential to advance their translational utility. Statistical harmonisation techniques such as ComBat^76,77^ can help mitigate site and scanner differences in multi-cohort applications, while normative modelling could establish reference distributions across older adults, enabling personalised assessments of deviation from typical brain health profiles. Such approaches will facilitate reproductivity, promote cross-study integration, and ultimately enable the use of these multimodal composites as scalable markers of brain health across diverse research and clinical contexts. In the interim, researchers can derive relative composites scores within their own datasets using the published factor loadings and cohort means (see section ***Statistical Analysis***) with acknowledgment that these scores may not be directly comparable between sites or cohorts.

### Study strengths and limitations

The present study included a range of high quality measures derived from multiple modalities that place emphasis on data quality and completeness through study inclusion and quality control procedures. Using agnostic, data-driven approaches, we identified and validated underlying constructs of brain health across modalities. A secondary strength of the confirmatory factor analysis approach is that for constructs to replicate in an external cohort, they cannot be driven primarily by confounding factors such as differences in MRI acquisitions and processing, or biobanking and biomarker assay protocols. The confirmatory factor analysis suggests that the validated constructs can withstand ‘batch effects’ resulting from cohort or methodological differences across studies. Equally, this may also explain why certain measures, such as HDL cholesterol, did not replicate into the confirmatory factor analysis, as their covariance structure may have been more sensitive to cohort- or method-specific influences. Nevertheless, future research should directly evaluate robustness to methodological variation (e.g., software pipelines, MRI acquisition parameters) by repeating confirmatory factor analysis with variables derived from alternative methods.

Despite these strengths, there are limitations to consider. First, the cross-sectional design limits interpretation of directionality between constructs and cognition. It remains unclear whether changes in the composite scores precede, follow, or co-evolve with cognitive decline. Second, while our cohorts are well-characterised, they may not capture the full demographic, vascular and pathological heterogeneity seen in the general population, which may be especially true for the smaller MCI and AD dementia samples. Applying these constructs in more diverse and clinically enriched cohorts will be critical for evaluating their translational utility. Lastly, a small proportion of the CN group were twins (15%). As twins share genetic and early environmental influences, their inclusion may introduce non-independence of observation^78,79^, potentially influencing covariance structure and parameter estimates. However, this study represented a validation of latent factors previously identified in a cohort without twins, reducing the likelihood that relatedness meaningfully influenced the observed factor structure. Nevertheless, future work should account for relatedness to further evaluate the robustness of biomarker covariance patterns and their associations with cognition and diagnostic status.

## Conclusion

In summary, we demonstrated a principled, multivariate framework for integrating multimodal biomarkers to characterise distinct domains of brain health. The confirmatory factor analysis approach strengthens confidence in this framework, demonstrating that empirically derived constructs can be reproduced across independent cohorts and reflect biologically meaningful processes. The identified composites are not disease-specific; they provide a means to probe underlying domains of brain health, reduce measurement noise inherent to individual markers, and integrate complementary information across modalities.

Importantly, the value of this work lies not only in the specific factors identified here, but in illustrating how complementary MRI, biofluid, and cardiovascular measures can be systematically combined to probe underlying dimensions of brain health. This framework is inherently adaptable and can be extended to incorporate emerging biomarkers and applied across populations and disease contexts. With further incorporation of data harmonisation and normative modelling approaches, this latent construct and composite framework holds extensive translational potential.

Future research should explore the application of this approach and composites in other neurological and systemic disease, evaluate utility for patient stratification in early intervention clinical trials, and examine capacity to track neurodegenerative progression. Collectively, these findings support the use of multivariate, multimodal integration as a flexible and generalisable strategy for advancing mechanistic understanding of cognitive decline and neurodegenerative disease.

## Data availability

The datasets generated and/or analysed during the current study are not publicly accessible but will be made available upon request. Due to ethics constraints, data will be shared on a project-specific basis. Depending on the nature of the data requested, evidence of local ethics approval may be required.

## Acknowledgements

The authors thank the Brain and Cognitive Health (BACH) cohort study and Prospective Imaging Study of Ageing (PISA) participants for their invaluable time and contribution to this research.

## Supplementary material

**eFigure 1.**
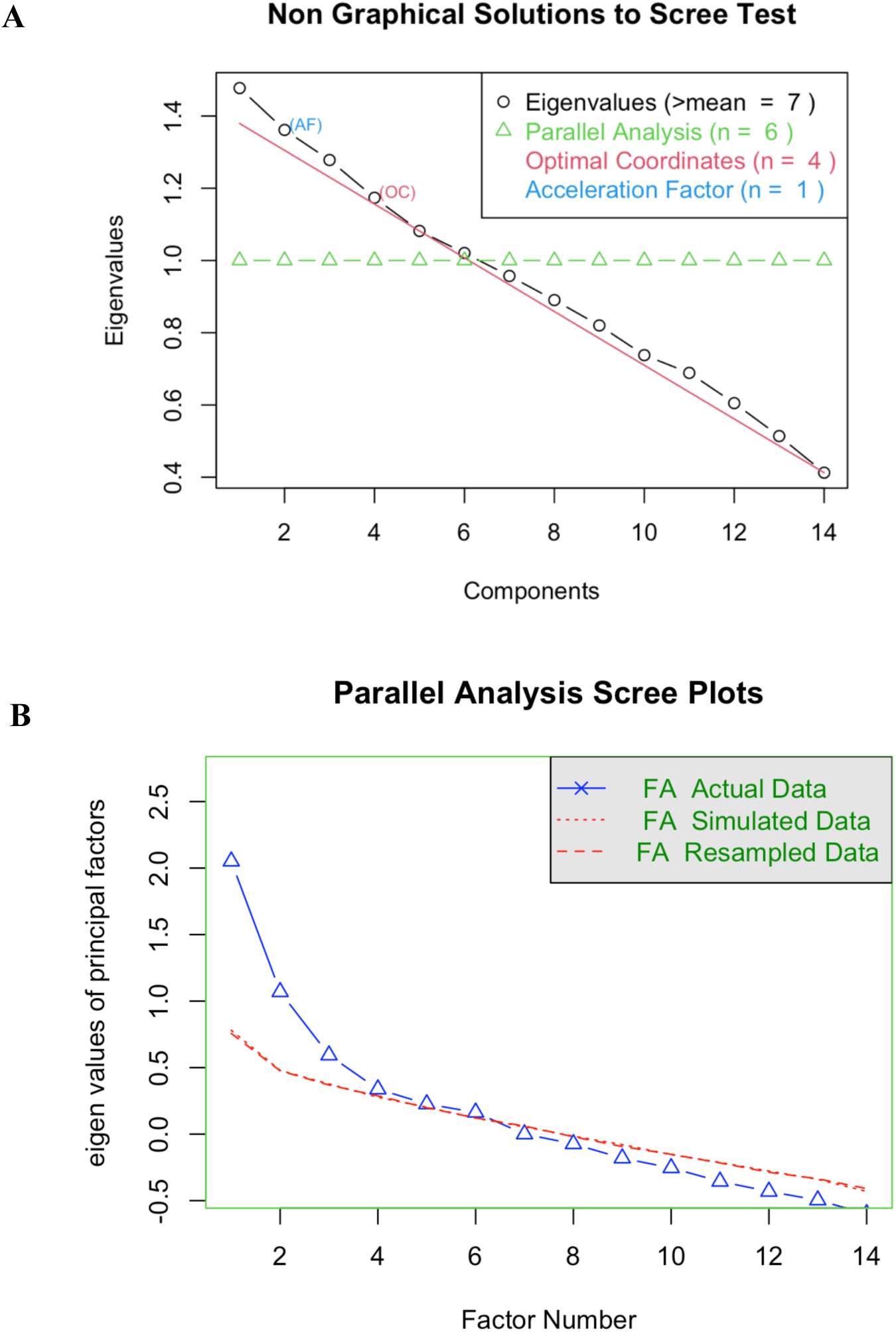
Exploratory Factor Analysis (EFA) Scree Plots. Scree plots were used to determine the numbers of factors to retain. **(A) Non-Graphical Solutions to the Scree Test:** This plot compares multiple factor retention methods. Parallel analysis (green) suggested 6 factors, optimal coordinates (red) suggested 4, and the acceleration factor method (blue) suggested 1. A four-factor solution was supported as a balance between these methods, theoretical interpretability and model fit indicies. **(B) Parallel Analysis Scree Plot:** This plot offers a robust refinement of the scree test findings. The retainment of n-factors was supported if their eigenvalues (blue) exceeded those of the simulated (dotted red) and resampled (dashed red) data. This method also showed support for a four-factor solution.

**eTable 1.** The five-factor model fit trialled in the exploratory factor analysis without pTau217.

|  | <u>Factor 1:</u> | <u>Factor 2:</u> | <u>Factor 3:</u> | <u>Factor 4:</u> | <u>Factor 5:</u> |
| --- | --- | --- | --- | --- | --- |
| **HDL Cholesterol** | **0.40** | -0.03 | 0.21 | 0.18 | 0.06 |
| **BMI** | **-0.41** | 0.14 | 0.05 | -0.25 | 0.05 |
| **Hippocampal Volume** | **0.82** | -0.07 | -0.06 | -0.03 | -0.01 |
| **Ventricle Volume** | **-0.70** | -0.05 | -0.11 | -0.01 | 0.03 |
| **CBF** | **0.41** | 0.12 | -0.03 | 0.02 | -0.06 |
| **BG ePVS** | **0.39** | **0.32** | 0.014 | -0.16 | 0.16 |
| **fwFA** | -0.08 | **0.68** | -0.07 | 0.20 | -0.05 |
| **Cortical Thickness** | 0.04 | **0.56** | 0.14 | -0.19 | 0.04 |
| **WM ePVS** | 0.04 | 0.02 | **0.99** | 0.02 | 0.00 |
| **Free Water** | -0.26 | -0.11 | **0.55** | 0.18 | -0.04 |
| **pTau181** | -0.05 | 0.14 | 0.30 | **0.30** | -0.19 |
| **GFAP** | 0.08 | 0.06 | 0.06 | **0.62** | -0.05 |
| **NfL** | -0.04 | 0.03 | -0.01 | **0.70** | 0.11 |
| **Aβ42/40** | -0.01 | -0.01 | -0.01 | 0.02 | **1.00** |

**Note:** loadings for each variable are reported, where variables with a loading of 0.30 or greater were considered to meaningfully contribute to the latent factor.
Aβ =amyloid-beta; BG=basal ganglia; BMI=body mass index; CBF=cerebral blood flow; ePVS=enlarged perivascular space; FW=Free Water; fwFA=Free Water-corrected fractional anisotropy; GFAP=glial fibrillary acidic protein; HDL= high-density lipoprotein; NfL=neurofilament light chain; pTau=phosphorylated tau; WM=white matter.

**eFigure 2.**
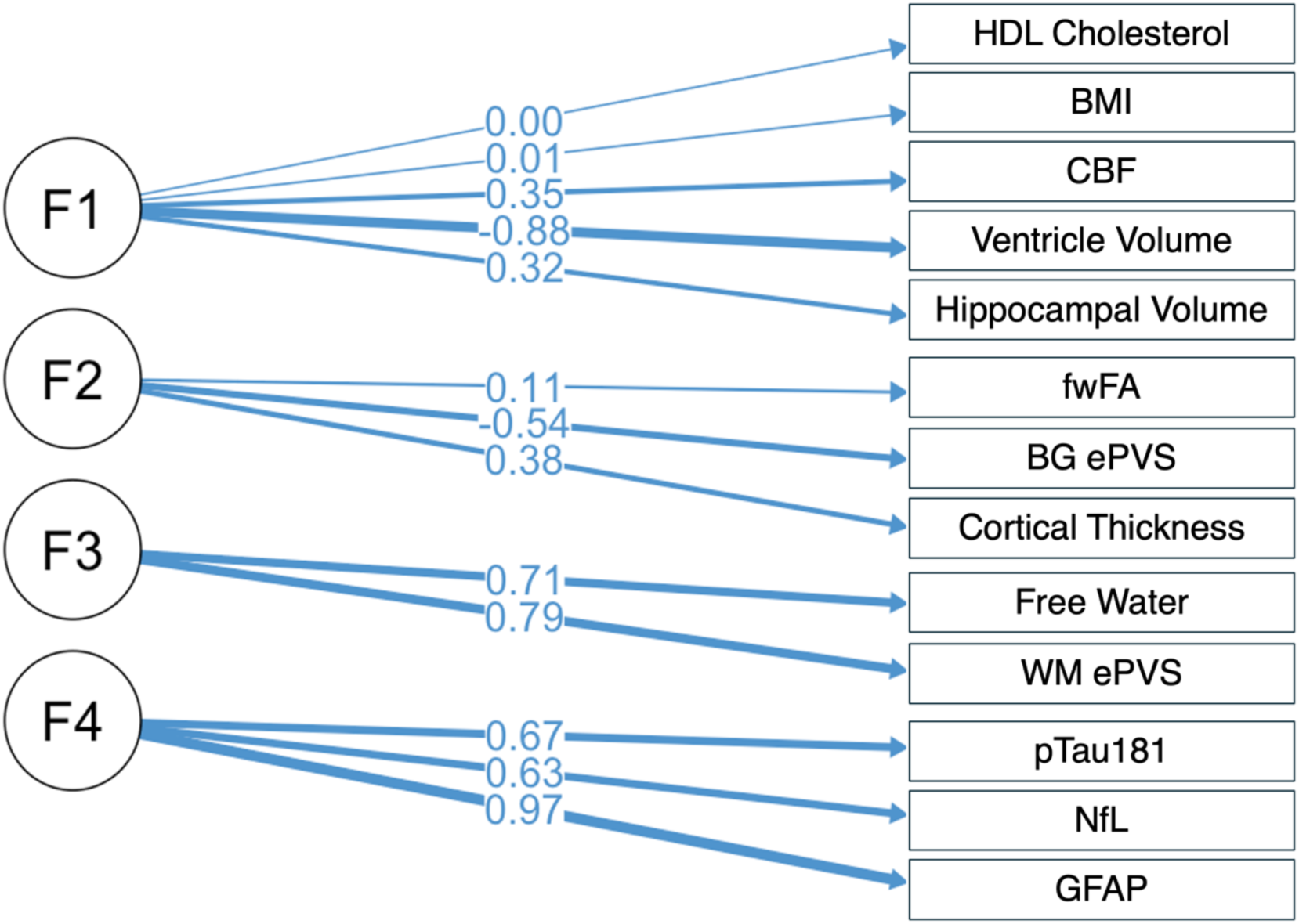
Unrefined Confirmatory Factor Analysis. Outcomes from the unrefined confirmatory factor analysis. Factor 1 (F1) “Brain & Vascular Health”, Factor 2 (F2) “Structural Integrity”, Factor 3 (F3) “WM Fluid Dysregulation”, Factor 4 (F4) “Blood Biomarkers”. Values represent variable standardised factor loadings onto the hypothesised exploratory factor analysis structure, with corresponding line width. The unrefined model demonstrated marginal fit (posterior predictive p-value = 0.002). BG=basal ganglia; BMI=body mass index; CBF=cerebral blood flow; ePVS=enlarged perivascular space volume; fwFA=Free Water-corrected fractional anisotropy; GFAP=glial fibrillary acidic protein; NfL=neurofilament light chain; pTau=phosphorylated tau; WM=white matter.

### Propensity-Matched Controls

To conduct analyses that account for the large sample size discrepancy between the cognitively normal health control (CN) group (n=157), and the MCI (n=18) and AD dementia (n=22) groups, we identified a propensity-matched subgroup of CN. Using the ‘matchit’ software in R (v4.7.2)^2^, for each MCI and AD dementia participant, a CN participant was selected based on a nearest neighbour approach for the combination of age and sex. The 40 matched CN participants were not significantly different from the MCI or AD groups in age, sex or education as tested by Wilcoxon t-test or Fisher’s Chi-Squared test (all p>.05; **eTable 2**).

**eTable 2.** PISA Cohort Summary: Propensity-Matched Controls.

|  | **Matched CN**<br>n=40 | **MCI**<br>n=18 | **AD**<br>n=22 |
| --- | --- | --- | --- |
| **Demographics** |  |  |  |
| Age at MRI, years | 65.71 (5.85) | 69.31 (6.89) | 65.47 (6.94) |
| Women (n, %) | 21 (52.5%) | 7 (38.9%) | 13 (59.1%) |
| Education, years | 13.68 (2.99) | 12.61 (3.62) | 14.18 (3.40) |
| *APOE* ε4 carrier (n, %) | 23 (57.5%) | 9 (50.0%) | 14 (63.6%) |
| **Cognition** |  |  |  |
| Similarities, total score<sup>^</sup> | 33.67 (4.51) | 26.94 (4.99) | 23.95 (8.94) |
| RAVLT-A, total score<sup>^</sup> | 13.22 (1.67) | 9.61 (3.09) | 11.00 (3.24) |
| Topographical Recognition, total score<sup>^</sup> | 26.02 (2.64) | 22.00 (4.99) | 17.55 (6.02) |
| FAS, total score<sup>^</sup> | 39.67 (12.33) | 35.83 (13.05) | 27.09 (9.76) |
| **Cardiovascular Risk Indices** |  |  |  |
| BMI, index<sup>^</sup> | 26.92 (4.24) | 26.40 (3.97) | 25.64 (3.93) |
| Systolic Blood Pressure, mmHg<sup>^</sup> | 139 (18) | 137 (18) | 141 (16) |
| HDL Cholesterol, mmol/L<sup>^</sup> | 1.58 (0.46) | 1.51 (0.47) | 1.70 (0.47) |
| LDL Cholesterol, mmol/L<sup>^</sup> | 3.00 (1.05) | 2.89 (1.05) | 3.56 (0.88) |
| **Neuroimaging** |  |  |  |
| WM Free Water, volume fraction | 0.087 (0.014) | 0.089 (0.014) | 0.084 (0.017) |
| BG ePVS, fraction of BG region | 0.0051 (0.0022) | 0.0046 (0.0018) | 0.0055 (0.0026) |
| WM ePVS, fraction of NAWM region | 0.0036 (0.0032) | 0.0026 (0.0022) | 0.0022 (0.0023) |
| GM cerebral blood flow, ml/100g/min | 21.86 (5.04) | 19.95 (4.75) | 18.43 (4.27) |
| WMH volume, mm<sup>3</sup> (median, IQR) | 373.5 (795.25) | 1017.5 (4437.25) | 561.5 (1438.5) |
| Cortical thickness, mm | 2.38 (0.07) | 2.30 (0.06) | 2.27 (0.09) |
| Ventricle volume, fraction of eTIV | 0.0148 (0.0047) | 0.0283 (0.0144) | 0.294 (0.0111) |
| Hippocampal volume, fraction of eTIV | 0.0056 (0.004) | 0.0046 (0.005) | 0.0048 (0.007) |
| fwFA, degree of anisotropy | 0.507 (0.011) | 0.508 (0.013) | 0.497 (0.035) |
| PET Amyloid Centiloid (median, IQR)^ | -4.96 (16.17) | 54.56 (70.05) | 99.14 (40.50) |
| PET amyloid centiloid <20 (n, %)^ | 6 (15.0%) | 12 (66.7%) | 22 (100%) |
| **Plasma biomarkers** |  |  |  |
| Fasted blood sample (n,%) | 25 (62.5%) | 5 (27.8%) | 3 (13.6%) |
| Aβ42/40 ratio | 0.068 (0.015) | 0.069 (0.011) | 0.064 (0.008) |
| GFAP, pg/mL | 80.39 (58.82) | 97.39 (45.88) | 170.81 (54.99) |
| NfL, pg/mL | 17.54 (13.05) | 18.44 (13.30) | 23.73 (7.87) |
| pTau181, pg/mL | 7.87 (6.38) | 20.53 (18.06) | 25.80 (20.37) |

**Note:** mean (SD) reported unless otherwise specified.
*APOE* ε4 carrier defined as having at least one ε4 allele (i.e. either heterozygote or homozygote).
^Some outcomes were not available for some participants:
PET Amyloid Centiloid: CN 38, MCI 18, AD 20.
BMI: CN 38, MCI 11, AD 11.
Systolic BP: CN 38, MCI 9, AD 10.
HDL and LDL Cholesterol: CN 38, MC 18, AD 2.1
No significant demographic differences in MCI or AD groups compared to the matched CN group (via Wilcoxon t-test or Fisher's Chi-Squared test (all $p > .05$ )).
Aβ=amyloid-beta; AD=Alzheimer's disease; *APOE*=apolipoprotein-E; BG=basal ganglia; BMI=body mass index; CBF=cerebral blood flow; CN=cognitively normal health control; ePVS=enlarged perivascular space volume; eTIV=estimated total intracranial volume; fwFA=Free Water-corrected fractional anisotropy; GFAP=glial fibrillary acidic protein; HDL=high-density lipoprotein; LDL=low-density lipoprotein; MCI=mild cognitive impairment; NAWM=normal appearing white matter; NfL=neurofilament light chain; PET=positron emission tomography; pTau=phosphorylated tau; RAVLT=Rey Auditory Verbal Learning Test; TMRT=Topographical Recognition Memory Test; WM=white matter; WMH=white matter hyperintensity volume.

### BG ePVS Post-Hoc Analysis

In the original factor analysis, the “Structural Integrity” construct was comprised of greater cortical thickness, higher Free Water-corrected fractional anisotropy (fwFA) and greater basal ganglia enlarged perivascular space volume (BG ePVS). However, in the present confirmatory factor analysis, higher cortical thickness loaded with *lower* BG ePVS, contrary to previous findings. To better understand this change of loading direction, we conducted a post-hoc linear regression investigating the relationship between BG ePVS and age, adjusting for sex and intracranial volume, using the same control sample as the CFA. We found that, in contrary to data in our original cohort, BG ePVS was *positively* associated with age (β=0.447, SE = 0.072, p<.001; **eFigure 3**), suggesting that this direction change is due to a change in the BG ePVS variable and not the construct itself.

**eFigure 3.**
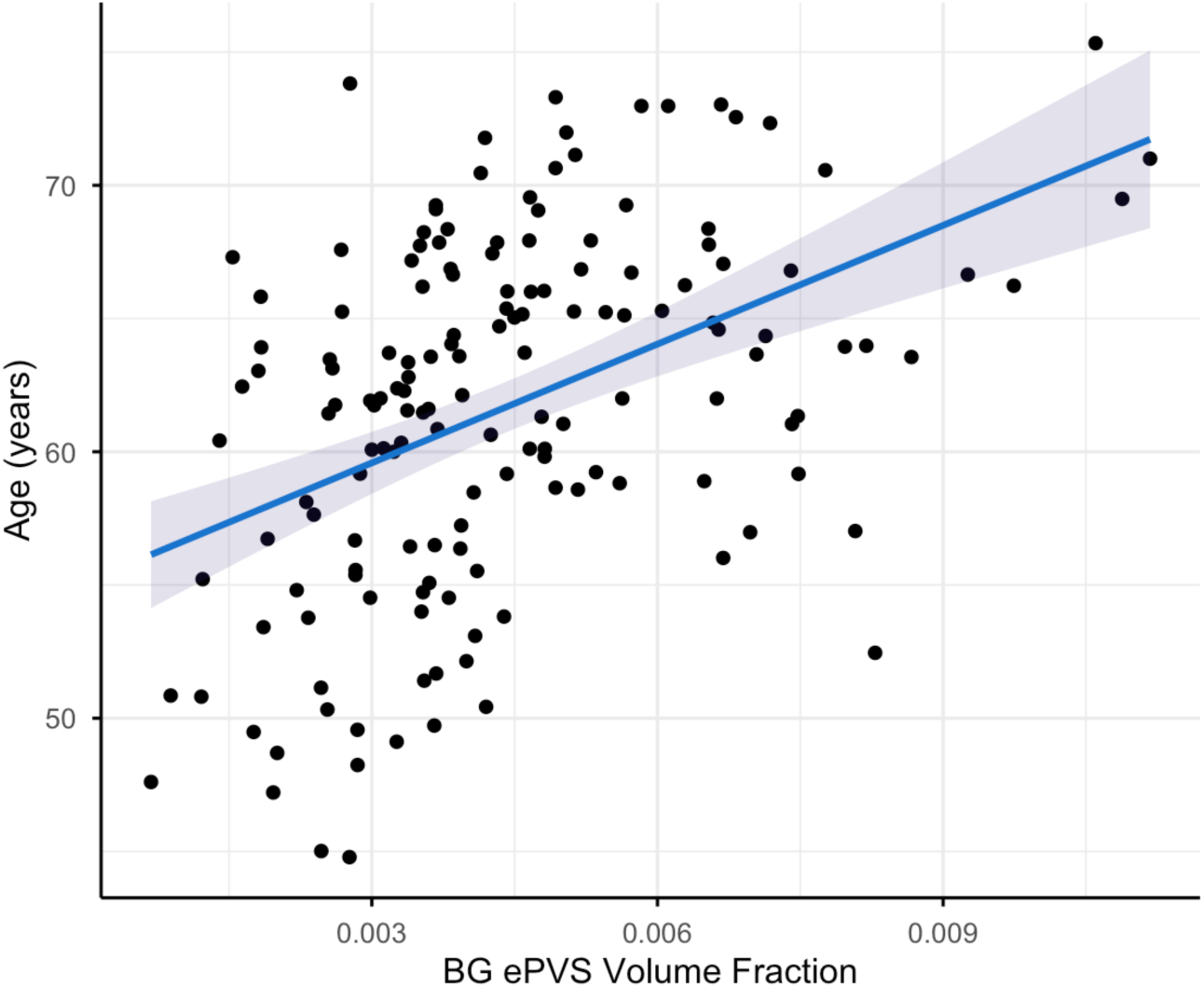
Positive Association between Age and BG ePVS in CN. Linear regression showed a positive association with age and basal ganglia enlarged perivascular space (BG ePVS) volume fraction in the cognitively normal controls (CN), which was contrary to the negative association found in the original EFA cohort.

**eFigure 4.**
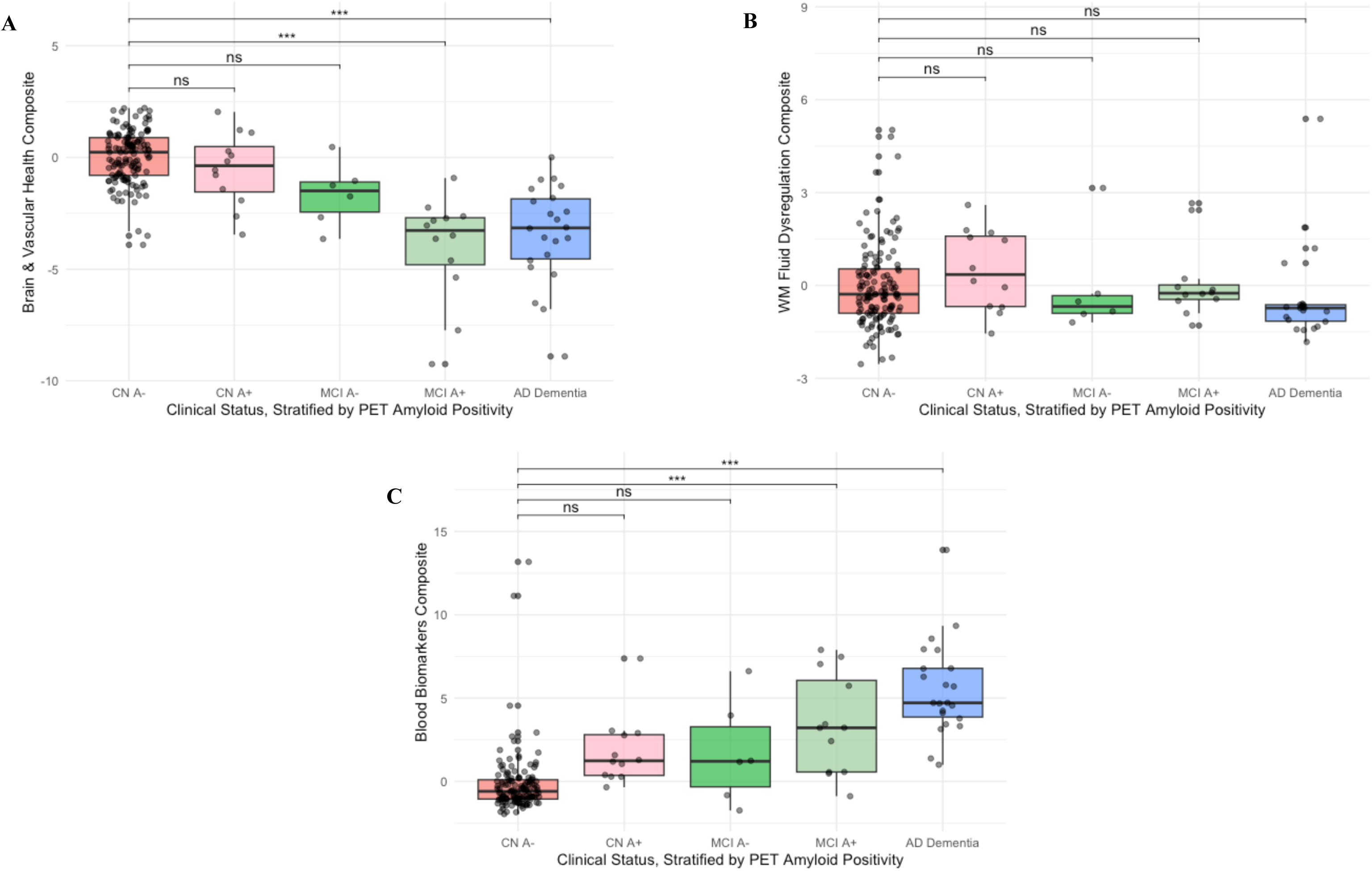
Composite score differences between CN, MCI and AD dementia groups, stratified by amyloid positivity. Differences across cognitively normal controls (CN), mild cognitive impairment (MCI) and Alzheimer’s disease (AD) dementia groups further stratified by PET amyloid positivity, reported for: (**A**) Brain & Vascular Health composite scores; (**B**) White Matter (WM) Fluid Dysregulation composite scores; (**C**) Blood Biomarkers composite scores. CN A- n=138, CN A+ n=12; MCI A- n=6, MCI A+ n=12; AD dementia (A+) n=22. A+=PET amyloid centiloid >20; A-=PET amyloid centiloid ≤20. *p<.05, **p<.01, ***p<.001, ns=did not meet statistical significance (p>.05).

**eFigure 5.**
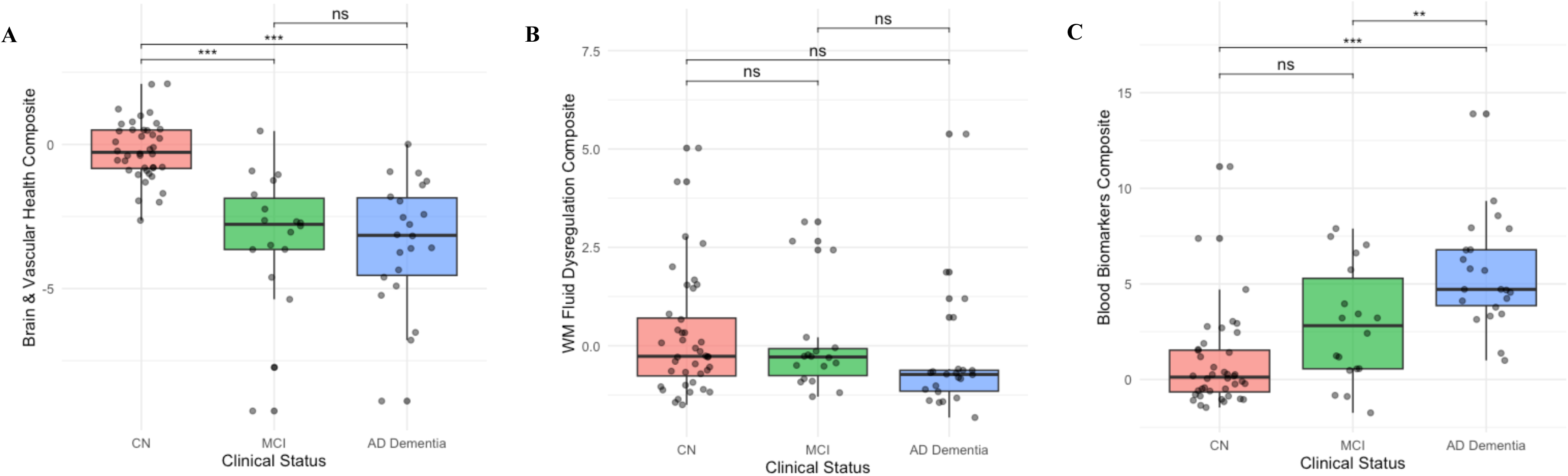
Composite score differences between propensity-matched CN, MCI and AD dementia groups. Differences across propensity-matched cognitively normal controls (CN), mild cognitive impairment (MCI) and Alzheimer’s disease (AD) dementia groups reported for: (**A**) Brain & Vascular Health composite scores; (**B**) White Matter (WM) Fluid Dysregulation composite scores; (**C**) Blood Biomarkers composite scores. *p<.05, **p<.01, ***p<.001, ns = did not meet statistical significance (p>.05).

